# Non-linear modelling of motor development in typically developing children and youth using robotic assessments

**DOI:** 10.1101/2023.11.01.23294040

**Authors:** Stephan C.D. Dobri, Stephen H. Scott, T. Claire Davies

## Abstract

**Background:** Clinical tasks are most often used to differentiate motor performance of individuals who have impaired function. However, these are not as accurate and repeatable as robotic tasks. Additionally, motor development occurs rapidly at early ages and slows as they reach adulthood, resulting in a non-linear model of performance. There is also evidence that variability in performance changes as children and youth age. Accurate normative models of performance are necessary to identify differentiate deficiencies in motor performance and to track the efficacy of therapies.

**Methods:** Two-hundred and eighty-eight participants who are typically developing (ages 5-18) completed a robotic point-to-point reaching task and an object hitting task using the Kinarm Exoskeleton. Exponential or quadratic curves were fit to performance parameters generated by Kinarm to model typical performance. These models included a linear term to account for changing variability with age.

**Results:** Most performance parameters showed improvement with age, and none showed deterioration. Some parameters showed large changes in variability in performance with age, up to a 74% decrease in the range of typical performance.

**Conclusions:** Reduced variability occurs with age, indicating the need to account for differences in variability when developing models of typical motor performance in children and youth. Models used to identify deficits in motor performance should account for changing variability in data and changing repeatability with age to increase accuracy of identification of deficits.

## Introduction

Motor development evolves in a non-linear manner such that rapid improvement occurs at the younger ages, and the rate of improvement slows until they have reached “near-adult” performance [1-3]. To be able to evaluate whether a child meets the typical milestones of motor performance (e.g. reaction time) their data can be plotted against age to generate curves that reflect developmental expectations of motor function. Clinical assessments rely on normative performance intervals to identify impairment on an individual level. For example, the Purdue Pegboard and Bruininks-Oseretsky Test of Motor Proficiency have been validated on large typically developing populations of typically developing children to determine normative performance intervals [4, 5]. These evaluations allow clinicians to test an individual and identify motor impairments.

Robotic assessments of motor function have been shown to be highly objective and repeatable; however, normative models of child development from robotic assessments have rarely been created are missing from current literature [6, 7]. Typically, researchers have used robotic assessments to make group comparisons between typically developing populations and clinical populations, such as children with cerebral palsy [7]. The purpose of this study was to create normative models of motor development using robotic assessments. These models of typical development can be used to identify deficits in performance in various patient populations and to track the efficacy of therapies. Therefore, a key first step in this process is the generation of accurate and reproducible curves of developmental behaviour.

## Methods

### Participants

Two-hundred and eighty-eight typically developing participants were recruited from the Kingston, Ontario, Canada area. Participants were between 5-18 years old. The cross-sectional study was approved by the Health Sciences and Affiliated Hospitals Research Ethics Board (HSREB) at Queen’s University, Kingston, Ontario, Canada (application number 6004951) in accordance with the Helsinki Declaration of 1975, as revised in 2000 and 2008. Participants were recruited through summer camps run by the university, word of mouth, and online flyers. Signed guardian consent and participant assent were obtained prior to testing. Participants were excluded if they had experienced recent concussion or had any physical or cognitive impairment that would affect completion of the study.

### Robotic Apparatus and Tasks

The Kinarm Exoskeleton Lab (Kinarm, Kingston, Ontario, Canada) was used in this study. Participants sat in a system-integrated modified wheelchair base with their arms supported against gravity. The robot allows free planar movement of the upper limbs while recording elbow and shoulder joint kinematics. Hand position and kinematics were calculated from the joint kinematics. Participants sat in front of a virtual reality display, with vision of their hands and arms occluded. Hand position feedback was displayed in each task on the virtual reality display. The robot has been described in detail previously [8]. Participants completed the Visually Guided Reaching (VGR) tasks and the Object Hit (OH) (Kinarm, Kingston, Ontario, Canada).

The Visually Guided Reaching (VGR) task assesses single handed goal-directed reaching [9]. There are four peripheral targets set in a square, 6 cm apart, with a fifth starting target in the centre of the square. Participants are instructed to reach as quickly and accurately as possible to each target as they appear. The peripheral targets appear in a pseudo-random order, six times each for a total of 24 reaches per arm. Each reach consists of moving out to the peripheral target, then back to the central target when it reappears. Participants complete the task once with both their dominant and non-dominant hands. This task is represented in Fig. 1A. In this case, the task is assessed using three different parameters Reaction Time (RT), Path-Length Ratio (PLR), and Min-Max Speed Difference (MMSD) [9]. Reaction Time (RT) is defined as the time (in seconds) from when the peripheral target appears to when hand movement onset occurs. Path-Length Ratio (PLR) refers to the ratio of the hand path travelled between movement onset and offset and the shortest path between two targets. Finally, the Min-Max Speed Difference (MMSD) relates to the difference between speed maxima and minima throughout the movement.

**Figure 1:**
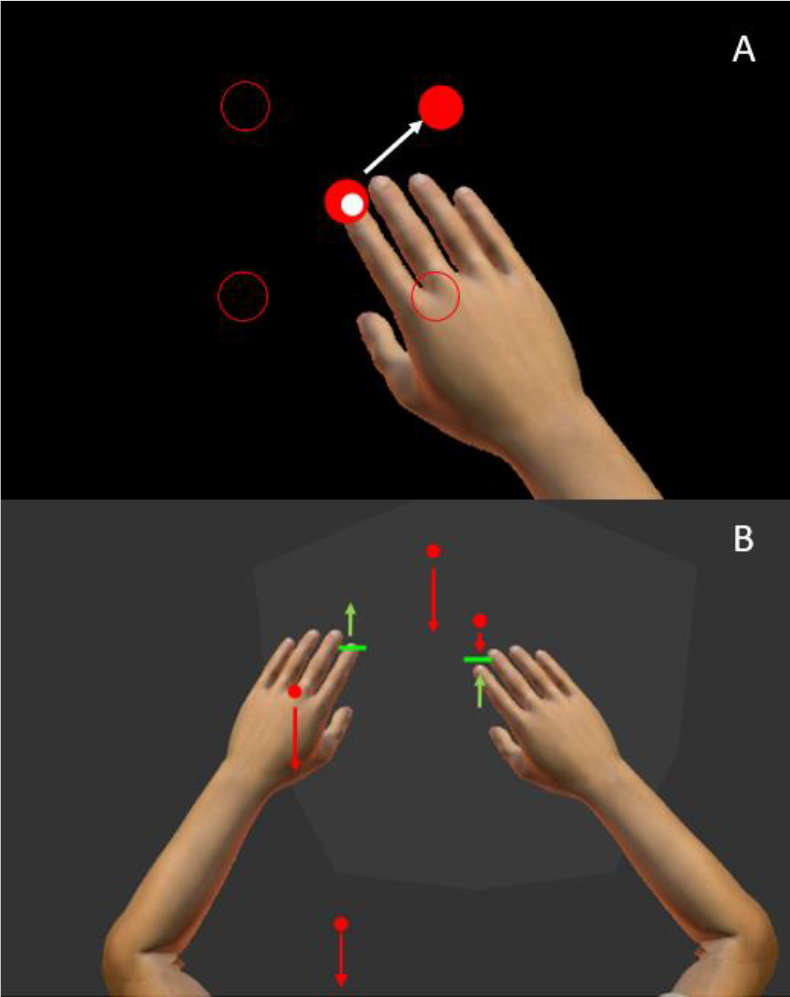
The Kinarm Exoskeleton Robot and representations of Visual Guided Reaching (VGR) and Object Hit (OH) tasks. A) VGR task in which the participant was waiting in the central target for a peripheral target to appear. The top-right target appeared so the participant must move the white dot representing hand position into the red target. The positions of the other peripheral targets are represented by the red circle outlines. B) a participant completing OH. The red balls are falling down the screen at different speeds, represented by the length of the red arrows.

The Object Hit (OH) task measures bimanual coordination [8, 10]. The task consists of red balls “falling” down the screen (from top to bottom) towards the participant, and the participant is instructed to hit the balls away from themselves. Both hands are used in this task and hand position feedback is provided by the visual representation of green paddles with which participants hit the balls. The robotic apparatus provides haptic and visual feedback when the ball is hit. The number of balls and speed at which they fall increases throughout the task (just over two minutes total time), increasing the difficulty as the task progresses. OH is represented in Fig. 1B. Three task parameters were chosen for assessment: Total Hits (TH), Hand Bias of Hits (HBH), and Movement Area Bias (MAB) [8, 10]. Total Hits (TH) is the total number of balls the participant hit with both hands of the 300 dropped down the screen. Hand Bias of Hits (HBH) describes a quantification of the bias in hits between the hands, calculated as: [(Hits Dominant – Hits Non-Dominant)/Total Hits]. The final parameter is the movement Area Bias (MAB) which is a quantification of the bias in the movement area between hands, calculated as: [(Movement Area Dominant – Movement Area Non-Dominant)/ (Movement Area Dominant + Movement Area Non-Dominant)]. Movement area is the area of the workspace (in m^2^) covered by a hand during the task, and it is determined by the convex hull (polygon) that includes the boundaries of the movement trajectory of the hand in question.

### Participant Data Analysis: Assessing Accuracy and Repeatability of Curve Fitting

Data for each parameter from both tasks were fit with one of two curves, exponential or quadratic, as shown below:

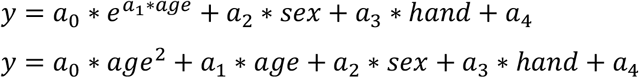

The variables “sex” and “hand” are dummy-coded variables (1 or 0) representing participant sex and the hand (dominant or non-dominant) that completed the task. The “hand” variable was not included in the model for the parameters from the OH task, as both hands were used during the task. The system does not allow for comparisons between right-hand versus left-hand dominance.

Curves were fit using the MATLAB function “nlinfit.m” (Mathwork, Inc., Massachusetts, USA). An exponential fit was used for parameters with only positive values. Exponential curves require a good initial estimate of the coefficients for the curve fitting function to produce accurate results [11]. Initial estimates were calculated for the coefficients a_0_, a_1_, and a_4_ in the exponential equation using a method described by Foss [11]. By taking the natural logarithm of the values and fitting a line, coefficients of the line were used as initial estimates for the exponential curve fitting. This method does not work with data that has negative or zero values as the logarithmic value would result in imaginary and infinite values. Thus, quadratic curves were fit to any parameter that included both positive and negative values.

Ten-fold cross-validation was used to avoid over-fitting models to the data. The data were divided into ten equal groups, nine groups were used to train the model and the tenth group was used to test the model. The fitting was completed ten times with each group being used once as the testing dataset. The model that performed best (according to the r-squared value of the test dataset) was then used as the model of the data.

Next, a linear model of the absolute value of the residuals relative to age was fit using the same ten-fold cross-validation technique. The linear model accounted for age-related changes in variability of the data. The model of residuals was used to calculate z-scores, as described by Altman [12]. An additional scaling factor was included to generate an unbiased measure of absolute deviation. The equations used to generate the z-scores are shown below:

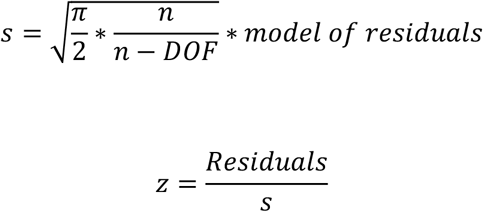

where DOF is the degrees of freedom of the model and n is the number of datapoints.

Three rounds of outlier removal were conducted on the calculated z-scores. The mean and standard deviation were calculated and any datapoints outside three standard deviations above or below the mean were identified as outliers. The mean and standard deviation were then re-calculated, excluding the outliers, and any new outliers were again identified. This process was completed to ensure that large outliers did not affect the subsequent testing for normality of the distribution of z-scores.

The z-scores were then tested for normality using the Shapiro-Wilks test, and the skew and kurtosis were calculated. If the z-scores failed the Shapiro-Wilks test but the absolute value of the skew was less than 0.6, and the kurtosis was between 2.4 and 3.6, the z-scores were considered “normal enough”, as outlined by Pearson and Please [13]. If the z-scores failed the Shapiro-Wilks test and the skew or kurtosis were outside the acceptable ranges, the data were transformed using the following transforms:

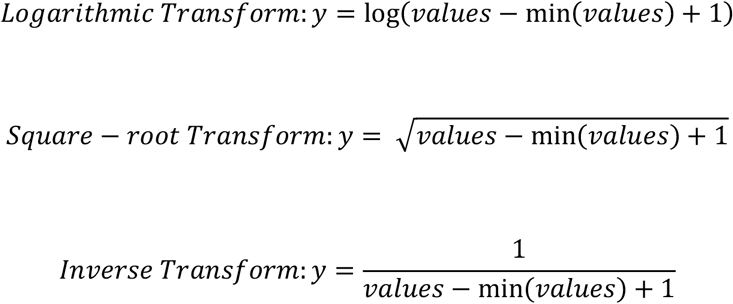

To ensure that the transforms calculated only real, finite numbers, each datapoint was shifted by subtracting the minimum value in the dataset and adding one. For example, if the minimum value of a parameter was three, each datapoint for that parameter would have the value 3 subtracted and then the value 1 added. The shifting ensured the new minimum value was one and all values were positive. The three transforms were applied separately to the data and the same curve fitting previously described was completed for the transformed data. The best transform produced normally distributed z-scores. If multiple transforms produced normal z-scores the transform with the highest r-squared value and a normal distribution was chosen. If all three transforms failed to produce normally distributed z-scores, the models from the untransformed data were kept for simplicity. The z-scores with a normal distribution were then used to create ranges of typical performance.

## Results

One can observe age-related differences in motor function from visual representations of performance parameters of reaching trials of the VGR task. Fig. 2 compares hand path trajectories and hand speed recordings from a participant in the youngest age group (5-10-years-old), and a participant from the oldest age group (13-18-years-old). Both participants were matched by sex and hand dominance, and the data are from reaches with their dominant hands. Individual lines represent separate reaching trials. Fig. 2A shows the hand path trajectories for reaching out and back. The younger participant (top) had more inter-reach variability than the older participant, as demonstrated by the wider spread of individual lines. They also had more corrective movements within each reach, as shown by the less-straight movements and overshooting of the targets (represented as black circles). These reaching behaviours were also quantified with the path-length ratio, and min-max speed difference.

**Figure 2:**
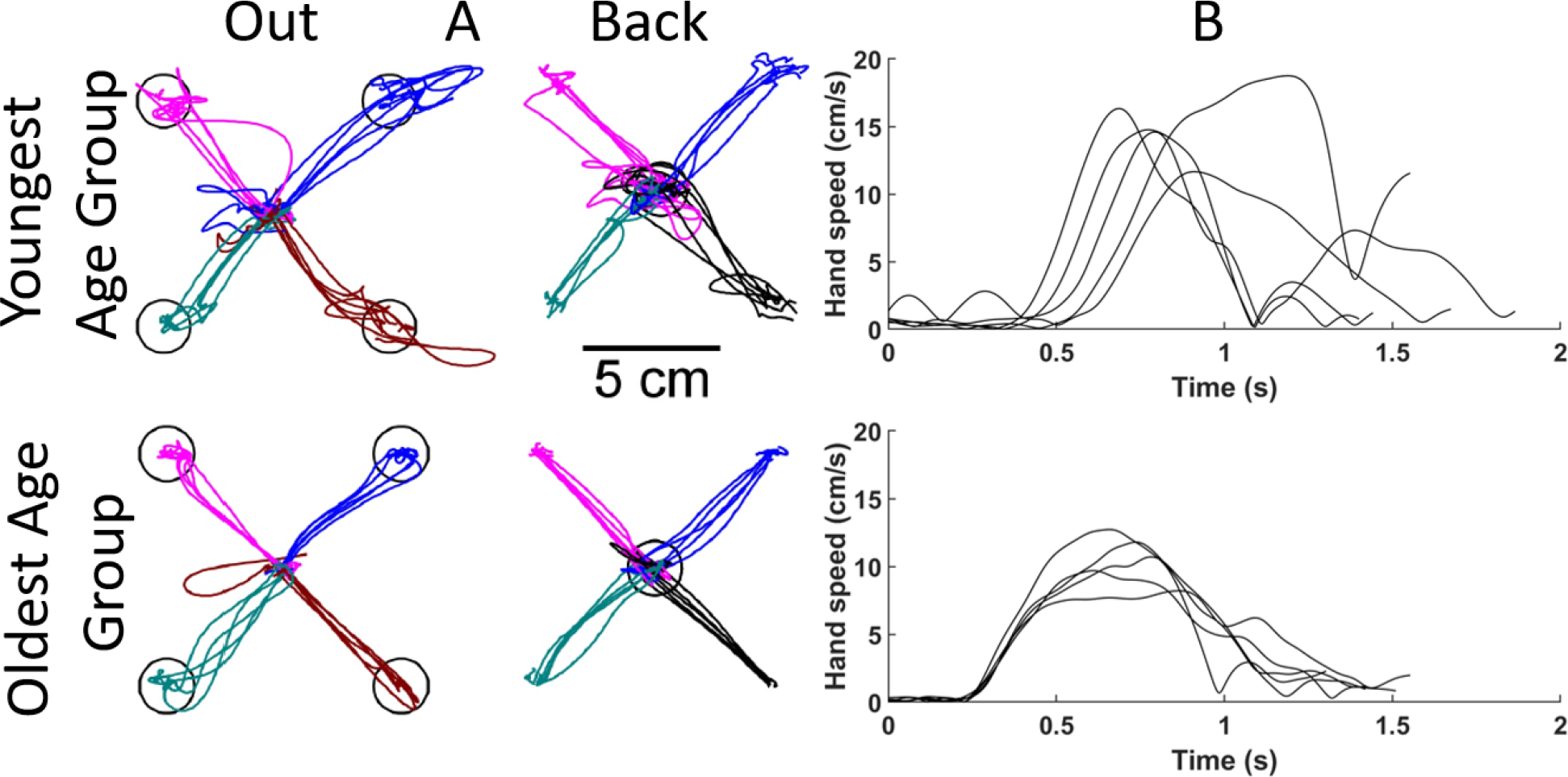
Comparisons of reaching for sex and handedness matched participants from the Visually Guided Reaching (VGR) task. The top row shows results from a younger participant (between 5- and 10-years-old), and the bottom from an older participant (between 13- and 18-years-old). A) Hand paths for each reach to each target. Each line represents an individual reaching trial. B) Hand speeds for reaches out to the bottom right target. Each line represents the hand speed during an individual reaching trial.

Fig. 2B shows hand speed profiles for different reaches. The younger participant had a slower reaction time, as seen by the later movement onset, and the speed profiles again show more corrective movements from the multiple speed peaks.

**Figure 2:**
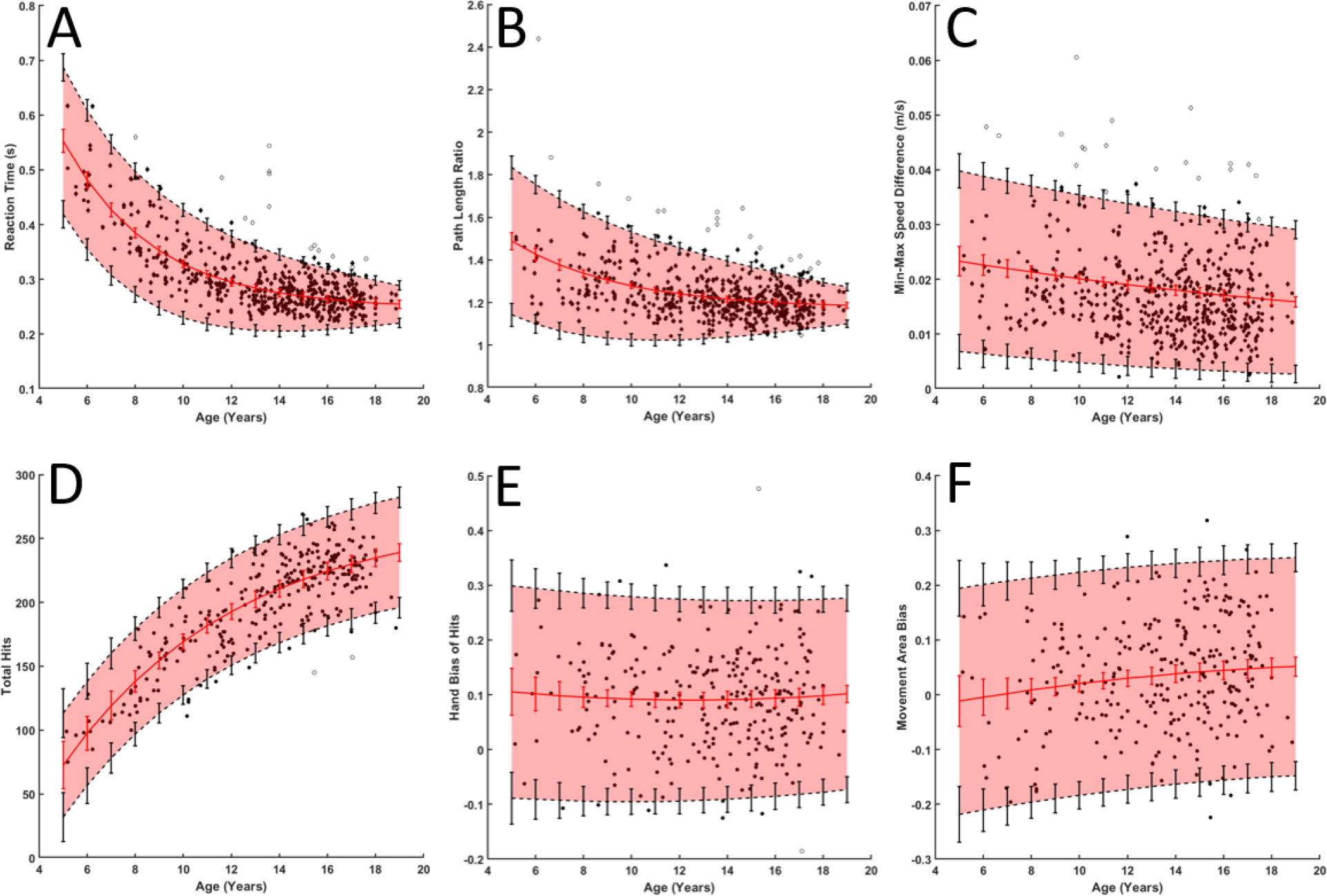
Six parameters from the two tasks with raw data and fitted curves. In each subplot, black dots denote individual participants, the red line and vertical red lines denote fitted curve and its 95% confidence interval, respectively. Black dashed lines and vertical lines denote 2.5 and 97.5% performance and associated vertical lines denote 95% confidence interval for each of these. The red shaded area is included to help visualize the central 95% range.

Curves were fit to the six performance parameters using the presented curve-fitting algorithm (Fig 3). The error bars were generated using the results from the simulations from uniform datasets discussed previously by Dobri et al. [14]. Fig. 3A-C show the three performance parameters from the VGR task, whereas Fig. 8D-F show the three parameters from the OH task. Both RT and PLR (Fig. 3A and B) show a decaying exponential relationship with age, and decreasing performance interval width. The typical performance interval widths (i.e. 95% band in shaded red) for RT decreased from 0.2680 s at age 5 to 0.0685 s at age 18, which reflects a 74% decrease in the variability in typical performance. Interval widths decreased from 0.6874 to 0.1867 for PLR (73% decrease). MMSD showed a linear relation with age, TH showed an increasing exponential relation, and HBH and MAB showed quadratic relations.

The performance parameters from VGR quantify some of the qualitative aspects of reaching movements noted in Fig. 2. The hand path traces showed that the younger participant deviated from a straight path more than the older participant. The deviation would result in a larger PLR, which would decrease as participants get older, as is seen in Fig. 3. As noted in the speed plots in Fig. 2, the reaction time of the older participant is shorter than the younger participant and this is also reflected in RT in Fig. 3. Table 1 in the Supplemental Material shows the curve fitting results for the six parameters highlighted in Fig. 3. The fit type (exponential or quadratic) is represented by an “E” or “Q” superscript beside the parameter name in the table. All parameters were normally distributed without the need for transformation to achieve a Gaussian distribution of the z-scores. The r-squared values for the fitted curve ranged from 0 to 0.831 and 0 to 0.239 for the fit to the absolute values of the residuals for these six parameters. The number of outliers identified ranged from zero (Movement Area Bias) to 18 (Path Length Ratio and Min-Max Speed Difference), meaning up to 3% of the data were identified as outliers for the performance parameters in Table 1. Tables 2 and 3 in the Supplementary Material summarize the curve fitting results for all parameters of the OH and VGR tasks. R-squared values ranged from 0 to 0.835 for curve fits, and 0 to 0.357 for the residuals.

## Discussion

Normative models were created for all performance parameters for both Visually Guided Reaching (VGR), and Object Hit (OH) using the described algorithm. The dataset of typically developing participants was nearly twice the size of the next largest study of robotic tasks undertaken by groups of typically developing children and youth (288 participants compared to 155) [7, 15-17]. The size of the dataset increased the repeatability of the curve fitting. Since handedness was represented by a dummy coded variable, data from both hands for all participants were used to generate normative ranges for VGR parameters, effectively doubling the dataset size to 576 datapoints for that method. Using the confidence intervals proposed by Dobri et al. from simulations, the confidence in the curves (in units of standard deviation) would be between 0.21 and 0.68 with 155 datapoints, 0.15 and 0.47 for 288 datapoints, and 0.10 and 0.32 for 576 datapoints [14]. The decreased size of the confidence intervals that result as a child reaches near-adult performance in the curves of typically developing participants better ensures that the model can accurately identify motor control deficits in data from new participants. The decreased confidence interval widths are most important for participants whose data is near the edges of the normative performance range. For example, if a z-score of within +/-2 indicated typical performance this would have to be expanded to +/-2.68 for portions of the curve generated from 155 datapoints to account for confidence in the curve fitting. This would decrease by 8% and 13% for 288 and 576 datapoints, respectively.

This is the first time that variable? confidence intervals have been included in normative models of motor development in children and youth generated from robotic assessments. Including the confidence intervals quantifies the reliability of the curve fitting and identification of motor function deficits. Reporting confidence intervals on statistical tests is common in other disciplines and should be more common for these types of normative models. It is important to include such information to allow other researchers to fully understand the strength of conclusions drawn from the results of analysis.

The authors have not found other work that created normative models that account for changing variability with age [7]. All other research used a fixed value for the width of the normative performance range across the ages assessed. The changing typical performance ranges increased by 6% of the original value for TH and decreased by 74% and 75% for RT and PLR, respectively. The dramatic changes for both RT and PLR indicate that it would be important to include changing variability to effectively differentiate typical from atypical performance with these parameters, whereas it may not be as important for TH. For example, a fixed performance interval for RT would only accurately identify deficits in that specific parameter for participants between 12- and 13-years-old (where the changing range is closest to the fixed value). At 5-years-old for RT, the changing performance interval is 64% wider than the fixed interval, and it is 49% smaller at 18-years-old. The algorithm would inaccurately identify participants who did not have impairment as having impairments at younger ages, and inaccurately identify impaired performance as typical for older ages.

Another research group has also developed normative models of typically developing performance for the VGR and OH tasks using the Kinarm [16, 17]. The OH task performed by that group included a slightly different Kinarm environment as the workspace was slightly smaller than the version used here. The VGR task was the same between the two groups. Comparisons were made between the parameters Total Hits (TH), Reaction Time (RT), and Path Length Ratio (PLR). Curves that were quadratic with age were fitted to data from 155 typically developing participants. Data were separated between dominant and non-dominant hand performance before generating curves, and constant widths were used for the normative performance ranges. The same research group has also developed normative models of typical performance for the VGR and OH tasks using the Kinarm, and are compared below [16, 17].

The dataset collected for this manuscript was nearly twice the size of the dataset used by Grohs et al. and Hawe et al. [16, 17]. The mean and standard deviation of the age of the participants in each dataset were similar, although the standard deviation was larger for the smaller dataset (4.5 years vs 3.2 years). While the distribution across the age range for the smaller dataset is unknown, the larger standard deviation indicates the ages are more evenly spread than in the larger dataset. It is currently unclear which of the two datasets would produce more repeatable results: the smaller dataset with more even distribution or the larger dataset with a skew in the data. A non-uniform dataset would produce less repeatable results where data are sparse, such as at the younger ages for the dataset collected for this manuscript. There would likely be a smaller, uniform dataset that would produce similar repeatability to a larger, non-uniform dataset.

The widths of the performance ranges calculated for this manuscript increased from 82 to 87 hits for TH, and decreased from 0.268 to 0.083 seconds for RT and 0.693 to 0.210 for PLR. The fixed width ranges found in Grohs et al. and Hawe et al. were 82 hits for TH, 0.2 seconds for RT, and 0.4 for PLR [16, 17]. The fixed values are within the changing interval ranges for all three of these parameters; however, the differences in the ranges for RT and PLR are important. The range calculated by the algorithm for RT is initially 34% larger than the fixed width range and ends 59% smaller. Similarly, for PLR the changing width starts 73% larger and ends 48% smaller. These differences in the widths of the normative performance intervals are important for identifying motor control deficits on an individual basis and should be taken into account.

While previous works did not generate separate typical performance curves for male and female participants, some did generate separate curves for dominant and non-dominant hands [16, 17]. Inter-sex differences in development have typically been found to be insignificant [18, 19], so it is justifiable to create only one curve. Most other work did not account for sex [7]; however, this work included the dummy coded sex variable. The addition of this variable gives the algorithm slightly more accurate differentiation between typical and atypical performance on the basis of sex. The improved differentiation would only affect participants who were near the boundaries of the performance intervals, as the sex difference is often small relative to the magnitude of the performance parameter.

The importance of separating dominant and non-dominant hand performance in the typically developing population is not clear. There is contrasting evidence suggesting that there may be significant differences in performance or that there may not [1, 20, 21]. In a similar manner to the sex variable, inclusion of the hand variable gives the algorithm slightly better differentiation between typical and atypical performance. Additionally, it allows easy comparisons between dominant and non-dominant hand performance for pathological populations. Separating the dominant and non-dominant hand is important for many different pathologies, including cerebral palsy and developmental coordination disorder. Previous works creating normative models have either only used data from the dominant hand [2, 3], or generated models for the dominant and non-dominant hand separately [15-17]. Since it is unclear if there are significant differences between dominant and non-dominant hand performance in typically developing populations, it is not clear whether it is necessary to generate separate curves for each hand. The advantage of using the dummy coded variable is that data from both hands are used to generate the curves, thereby doubling the number of datapoints than if each hand were used separately. This doubling increases the repeatability of the curve fitting. The main drawback is the underlying assumption that both hands develop in the same manner. However, this is supported by results indicating no significant difference in performance between the hands [1, 20, 21]. Future work could compare the efficacy of both methods: separating data and generating different curves or combining data and including the dummy coded hand variable.

## Limitations

The main limitation of this work is the distribution of participant ages. While the participant group is large, there is a skew toward the older ages and nearly twice as many male participants as female. These skews are likely due to a recruitment bias. The most successful method of recruitment was partnering with an engineering summer camp for children and teens from 11- to 17-years-old. This skew resulted in poorer confidence intervals on the curve fitting when compared to a uniform dataset. As such, the authors would not recommend using these curves to identify motor control deficits in children under 9-years-old. Future work should involve recruiting more younger, and more female typically developing participants.

The curve fitting algorithm was given a very good initial estimate of the coefficients used to generate the data (within 1% of each coefficient), and the accuracy and repeatability were not compared for different ranges of input coefficients. Poor initial estimates would most likely result in poorer fits to the data. The issue of poorer fits can be partially fixed by changing parts of the curve fitting function, such as the number of optimization iterations or optimization parameters. These additional solutions have not been explored in this work.

### Clinical Implications

The normative models provided could be used by clinicians to identify impairment in certain parameters for any Kinarm users, as well as to target and track therapies. The models can be used for robotic assessments in the same way typical performance intervals are used in existing clinical assessments, such as the Purdue Pegboard and Bruininks-Oseretsky Test of Motor Proficiency [4, 5]. Clinicians could use the models to identify if a person has motor control deficits relative to others of the same age and sex. Therapies could then be targeted to the specific needs of the individual. These normative models could also be used to track the efficacy of therapies as they can record small improvements and large milestones, such as crossing the threshold into the normative ranges for different task parameters. The algorithm could be applied to other robotic assessments, with the Kinarm or other devices, to create normative ranges for measures of other aspects of function, such as position sense.

## Conclusions

The normative models provided could be used by clinicians to identify impairment in certain parameters for any Kinarm users, as well as to target and track therapies. The models can be used for robotic assessments in the same way typical performance intervals are used in existing clinical assessments, such as the Purdue Pegboard and Bruininks-Oseretsky Test of Motor Proficiency. Clinicians could use the models to identify if a person has motor control deficits relative to others of the same age and sex. Therapies could then be targeted to the specific needs of the individual. These normative models could also be used to track the efficacy of therapies as they can record small improvements and large milestones, such as crossing the threshold into the normative ranges for different task parameters. The algorithm could be applied to other robotic assessments, with the Kinarm or other devices, to create normative ranges for measures of other aspects of function, such as position sense.

## Data Availability

All data produced in the present study are available upon reasonable request to the authors.

## Notes

### Competing Interest Statement

S.H. Scott is the Chief Scientific Officer and co-founder of Kinarm, the company that develops the robot and software used in this article.

### Funding Statement

This work was supported by the Natural Sciences and Engineering Research Council under Grants 513272-17 and RGPIN-2016-04669, and an Ontario Research Foundation - Research Excellence grant (RE09-112).

### Author Declarations

The cross-sectional study was approved by the Health Sciences and Affiliated Hospitals Research Ethics Board (HSREB) at Queen's University, Kingston, Ontario, Canada (application number 6004951) in accordance with the Helsinki Declaration of 1975, as revised in 2000 and 2008. Signed guardian consent and participant assent were obtained prior to testing.

