## Supplemental Tables 2 and 3. for "Non-linear modelling of motor development in typically developing children and youth using robotic assessments"

### Supplementary Material

This document contains supplementary material for the article, “Non-linear modelling of motor development in typically developing children and youth using robotic assessments”. The document contains tables of the results of the curve fitting for all parameters that were fit from the Visually Guided Reaching and Object Hit Kinarm Standard Tasks.

*Table 1: Curve fitting results for the six highlighted performance parameters.*

| Parameter Names | Curve Fitting to Data | | | | | | Curve Fitting to Residuals | | | Measures of Normality | |
|---|---|---|---|---|---|---|---|---|---|---|---|
| | $a_0$ | $a_1$ | $a_2$ | $a_3$ | $a_4$ | R- squared | m | b | R-squared | Skew | Kurtosis |
| Reaction Time[E] | 1.152 | -0.265 | -0.003 | -0.009 | 0.260 | 0.774 | -0.003 | 0.068 | 0.239 | 0.552 | 2.945 |
| Path Length Ratio[E] | 0.935 | -0.216 | 0.046 | -0.038 | 1.162 | 0.447 | -0.007 | 0.175 | 0.176 | 0.502 | 3.094 |
| Min-Max Speed Difference[E] | 0.020 | -0.045 | 0.003 | 0.00 | 0.004 | 0.168 | 0.000 | 0.007 | 0.013 | 0.587 | 3.040 |
| Target Hits Total[E] | -386.146 | -0.136 | 7.726 | -- | 260.598 | 0.831 | 0.072 | 15.801 | 0.007 | -0.265 | 2.894 |
| Hand Bias of Hits[Q] | 0.000 | -0.007 | 0.002 | -- | 0.130 | 0.000 | -0.001 | 0.080 | 0.002 | 0.063 | 2.812 |
| Movement Area Bias[Q] | 0.000 | 0.010 | -0.006 | -- | -0.049 | 0.075 | 0.000 | 0.083 | 0.000 | -0.040 | 2.538 |

*Table 2: Curve fitting results for the Object Hit task.*

| Parameter Names | Curve Fitting to Data | | | | | | Curve Fitting to Residuals | | | Measures of Normality | |
|---|---|---|---|---|---|---|---|---|---|---|---|
| | $a_0$ | $a_1$ | $a_2$ | $a_3$ | $a_4$ | R- squared | m | b | R-squared | Skew | Kurtosis |
| Target Hits Total[E] | -386.146 | -0.136 | 7.726 | -- | 260.598 | 0.831 | 0.072 | 15.801 | 0.007 | -0.265 | 2.894 |
| Median Error[E] | -37.217 | -0.132 | 0.160 | -- | 72.959 | 0.695 | 0.111 | 1.527 | 0.143 | 0.021 | 2.594 |
| Miss Bias[Q] | 0.000 | 0.002 | 0.003 | -- | -0.008 | 0.067 | 0.001 | 0.009 | 0.039 | 0.048 | 2.720 |
| Hand Bias of Hits[Q] | 0.000 | -0.007 | 0.002 | -- | 0.130 | 0.000 | -0.001 | 0.080 | 0.002 | 0.063 | 2.812 |

| | | | | | | | | | | | |
|---|---|---|---|---|---|---|---|---|---|---|---|
| Targets Hit (Dominant Hand)$^{E}$ | -195.657 | -0.121 | 5.301 | -- | 145.237 | 0.777 | 0.239 | 8.251 | 0.051 | -0.220 | 2.871 |
| Targets Hit (Non-Dominant Hand)$^{E}$ | -173.384 | -0.142 | 3.864 | -- | 115.481 | 0.732 | 0.248 | 7.635 | 0.016 | 0.047 | 2.534 |
| Hand Speed (Dominant Hand)$^{E}$ | 0.195 | -0.010 | 0.019 | -- | 0.028 | 0.121 | -0.002 | 0.070 | 0.120 | 0.443 | 3.298 |
| Hand Speed (Non-Dominant Hand)$^{E}$ | 2.336 | 0.000 | 0.013 | -- | -2.142 | 0.073 | -0.002 | 0.063 | 0.123 | 0.387 | 3.079 |
| Hand Speed Bias$^{Q}$ | 0.001 | -0.017 | 0.008 | -- | 0.151 | 0.069 | 0.000 | 0.068 | 0.014 | -0.089 | 2.888 |
| Movement Area (Dominant Hand)$^{E}$ | -0.222 | -0.414 | 0.013 | -- | 0.089 | 0.184 | 0.000 | 0.018 | 0.006 | 0.091 | 2.443 |
| Movement Area (Non-Dominant Hand)$^{E}$ | -0.072 | 0.001 | 0.012 | -- | 0.154 | 0.135 | 0.000 | 0.021 | 0.013 | 0.157 | 2.410 |
| Movement Area Bias$^{Q}$ | 0.000 | 0.010 | -0.006 | -- | -0.049 | 0.075 | 0.000 | 0.083 | 0.000 | -0.040 | 2.538 |

*Table 3: Curve fitting results for the Visually Guided Reaching task.*

| | Curve Fitting to Data | | | | | | Curve Fitting to Residuals | | | Measures of Normality |
|---|---|---|---|---|---|---|---|---|---|---|

| Parameter Names | $a_0$ | $a_1$ | $a_2$ | $a_3$ | $a_4$ | R- squared | m | b | R-squared | Skew | Kurtosis |
|---|---|---|---|---|---|---|---|---|---|---|---|
| Posture Speed$^{E}$* | 0.133 | -0.001 | 0.000 | 0.000 | -0.127 | 0.130 | 0.000 | 0.003 | 0.113 | 0.748 | 3.218 |
| Reaction Time$^{E}$ | 1.152 | -0.265 | -0.003 | -0.009 | 0.260 | 0.774 | -0.003 | 0.068 | 0.239 | 0.552 | 2.945 |
| Initial Direction Angle$^{E}$* | 0.106 | -0.153 | 0.008 | -0.003 | 0.045 | 0.248 | -0.001 | 0.031 | 0.142 | 0.683 | 3.124 |
| Initial Distance Ration$^{E}$ | -0.284 | -0.100 | -0.004 | 0.002 | 0.941 | 0.235 | -0.003 | 0.094 | 0.070 | 0.133 | 3.054 |
| Initial Speed Ratio$^{E}$* | -0.164 | -0.244 | 0.000 | -0.001 | 0.989 | 0.227 | -0.002 | 0.041 | 0.155 | -1.034 | 3.113 |
| Speed Maxima Count$^{E}$ | 11.721 | -0.641 | -0.021 | 0.010 | 2.179 | 0.072 | -0.004 | 0.334 | 0.016 | 0.327 | 3.077 |
| Movement Time$^{E}$ | 1.364 | -0.284 | -0.062 | -0.038 | 0.900 | 0.268 | -0.001 | 0.121 | 0.006 | 0.391 | 3.190 |
| Path Length Ratio$^{E}$ | 0.935 | -0.216 | 0.046 | -0.038 | 1.162 | 0.447 | -0.007 | 0.175 | 0.176 | 0.502 | 3.094 |
| Max Speed$^{E}$ | -0.103 | -0.178 | 0.021 | 0.001 | 0.176 | 0.185 | 0.001 | 0.024 | 0.037 | 0.448 | 2.868 |
| Min-Max Speed Difference$^{E}$ | 0.020 | -0.045 | 0.003 | 0.00 | 0.004 | 0.168 | 0.000 | 0.007 | 0.013 | 0.587 | 3.040 |

Note that * indicates no transform created a Gaussian distribution of the residuals so the initial fit was used for simplicity.
